# Clinical Predictors of Outcome in Nonsegmental Vitiligo: A Prospective Cohort Study

**DOI:** 10.64898/2026.04.29.26352012

**Authors:** Lipsa Kumari, Shreya K Gowda, Sunil Nagpal, Vishal Gupta, Shivam Pandey, Kanika Sahni, M Ramam, Somesh Gupta

## Abstract

**Background:** Non-segmental vitiligo(NSV) shows marked heterogeneity in activity, progression, and treatment response. Reliable clinical markers that predict prognosis and patient-reported outcomes are lacking.

**Objectives:** To identify clinicodemographic and clinical predictors of disease extent, progression, repigmentation, treatment dependency, noticeability, and psychosocial impact in NSV.

**Methods:** In this prospective cohort study, 275 patients with NSV were followed for 12 months. Sixteen baseline variables, including demographic features, autoimmune history, and clinical markers (koebnerization, confetti and trichrome patterns, leukotrichia, mucosal, acral, and periorificial involvement), were recorded. Outcomes included body surface area(BSA), progression, repigmentation, treatment dependency, Vitiligo Noticeability Scale(VNS), and quality-of-life indices(VIS-22, DLQI, C-DLQI, F-VIS). Multivariable analyses and cluster analysis were performed at 6 and 12 months.

**Results:** Markers of disease activity leukotrichia, trichrome and confetti lesions, koebnerization, and mucosal, acral, and periorificial involvement were strongly associated with greater BSA, poor repigmentation, higher noticeability, and treatment dependency. Leukotrichia was consistent predictor of poor repigmentation and high VNS. Family history of autoimmunity predicted progression and treatment dependency. Early-onset vitiligo showed lower disease extent but greater family-related psychosocial burden. Cluster analysis identified severe, intermediate, and mild phenotypes with distinct therapeutic responses.

**Conclusions:** Simple clinical markers can stratify NSV patients into prognostic subgroups, enabling individualized treatment and counseling.

**Plain Language Summary:** Vitiligo behave variably in different people, some people may have slow-spreading course, while others develop widespread or persistent patches. In this study, we followed 275 people with non-segmental vitiligo for one year to find signs on the skin that could predict how the disease would behave and how it would affect daily life.

We found that features such as white hair within patches (leukotrichia), speckled (confetti) or three-colored lesions (trichrome), new patches appearing after injury (koebnerization), and involvement of the lips, mouth, hands, feet were linked to more severe disease, poorer response to treatment, and greater cosmetic concern.

A family history of autoimmune disease increased the risk of worsening vitiligo. Patients who developed vitiligo early in life had less skin involvement but greater emotional and family-related impact. These easily recognized signs can help doctors and patients plan treatment and set realistic expectations.

**Significance of the study:** Non-segmental vitiligo (NSV) has a heterogeneous and unpredictable clinical course with variable progression and response to therapy. However, robust prospective data linking these markers with long-term outcomes and patient-reported measures remain limited. In our prospective cohort of 275 patients, clinical markers such as leukotrichia, trichrome and confetti lesions, koebnerization, and acral/mucosal/periorificial involvement, were strongly associated with greater disease extent, poorer repigmentation, higher treatment dependency, and increased noticeability. Leukotrichia consistently predicted poor repigmentation. Thereby, prognostic stratification can also improve patient counselling regarding expected repigmentation, treatment duration, and psychosocial burden.

## Introduction

Vitiligo is an acquired depigmenting disorder that affects 0.36% of the general population, 0.67% of the adult population, and 0.24% of children.^1^ It is characterized by progressive loss of melanocytes, leading to the formation of milky white, sharply demarcated macules to patches. It affects all races with an almost equal male-to-female ratio and can begin at any age.

The course of vitiligo can be predicted fairly confidently for segmental disease. However, non-segmental vitiligo (NSV) behaves differently in different individuals, and currently there exists no method by use of which we can predict its course in a given person. Different authors have used different definitions for active vitiligo.

According to a review by Van Geel et al., the primary clinical indicators of active vitiligo are Koebner’s phenomenon, confetti-like depigmentation, and tri- or hypochromic lesions with indistinct borders.^1,2^ Clinical markers can serve as prognosticators and predictors of its course and future severity, and can guide appropriate counselling and threshold of starting treatment in patients.^3^ And prognostication at the outset can help in stratifying the patients which can guide the aggressiveness of treatment to be started and duration for which it should be continued. Thereby, we conducted a single-center, large prospective cohort study to determine the prognostic factors.

## Materials and methodology

This was a single-center, prospective cohort study conducted at a tertiary care center from July 2023 to June 2025, following approval by the institutional ethics board [IECPG-529-20/9/2023]. Consented patients of non-segmental or mixed vitiligo diagnosed clinically and/or by Woods lamp of any age and gender, and willing to return for follow-up, were included. Of 283 patients screened, 275 were enrolled after exclusion, 243 and 218 patients completed the 6-month and 12-month assessment (Figure 1). A detailed history of the patient was taken with clinical examination. Clinical photographs were taken for record and comparison during follow-up. A specific note of 16 factors was done that included such as age of onset, duration of disease, delay in initiation of treatment, history of any auto-immune condition, family history of vitiligo and other autoimmune diseases, presence of halo nevi, acral, mucosal, and periorificial involvement, trichrome pattern, confetti like lesions, Koebner’s phenomenon, leukotrichia, topical/phototherapy/systemic treatment given to the patients were administered at the discretion of the treating physician, without any modification by the primary investigator and BSA was calculated by Lund and Browder chart. Treatment were categorized into topicals only, topicals plus oral immunosuppressants, and topicals with oral immunosuppressants with phototherapy. Psychosocial impact was assessed based on the VISS-22 Score, and DLQI for adults ≥16years; or C-DLQI, and F-VIS for children <16 years and family members of affected children respectively.

**Figure 1.**
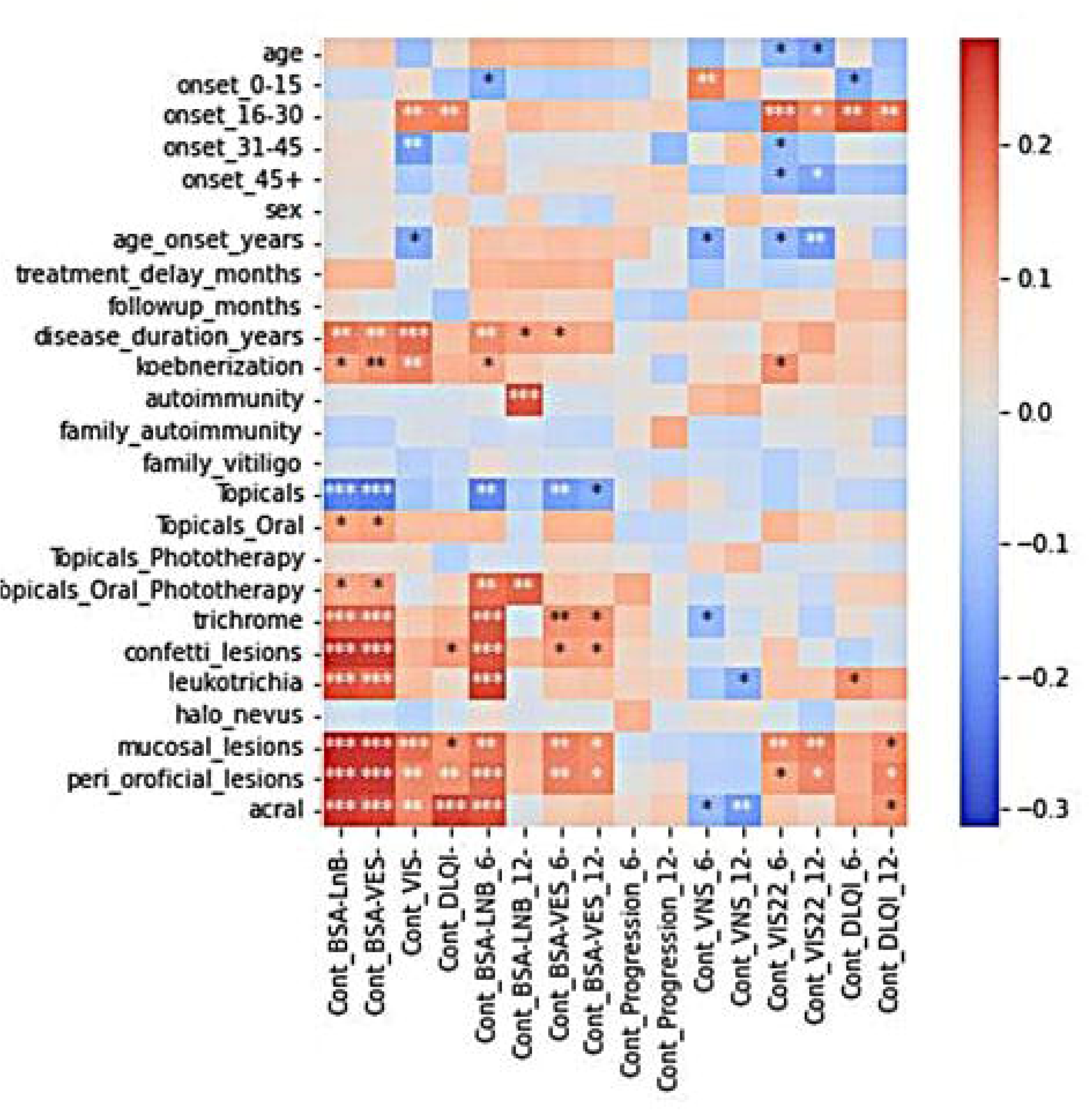
The methodology of the study (CONSORT flowchart).

Follow up at 6 and 12 months with a specific note of the following outcomes was made at 6 months and 12 months, such as progression (increase in size of lesions or new lesions), large extent of disease (≥5%body surface area), patient satisfaction level measured by Vitiligo Noticeability scale.^2^ Percentage repigmentation in overall patches was obtained by Investigator global assessment (IGA) and psychosocial impact was assessed based on the VISS-22 Score and DLQI; or C-DLQI and F-VIS for children <16 years at 6 and 12 months. All outcome measures were assessed in person by the investigator during baseline and follow-up visits.

The objective was to identify factors that affect the outcomes in non-segmental vitiligo in terms of progression, extent, repigmentation, treatment dependency, patient satisfaction and psychosocial impact.

## Results

We evaluated 275 patients with vitiligo to define clinicodemographic patterns and identify predictors of disease severity, progression, repigmentation, treatment dependence, noticeability, and psychosocial burden (Table 1). Collinearity analysis demonstrated significant interrelationships among clinical variables (Figure 2). Koebnerization showed strong positive associations with both trichrome and confetti patterns, which were also highly correlated. Mucosal, peri-orificial, and acral involvement exhibited mutual positive correlations. Treatment modalities were inversely related, with topical monotherapy negatively correlated with combination regimens that included systemic agents, phototherapy, or both. Combination therapy was more frequently used in patients with high-risk clinical markers such as koebnerization, confetti lesions, and trichrome patterns. Family history of vitiligo correlated positively with a family history of autoimmune disease

**Figure 2:**
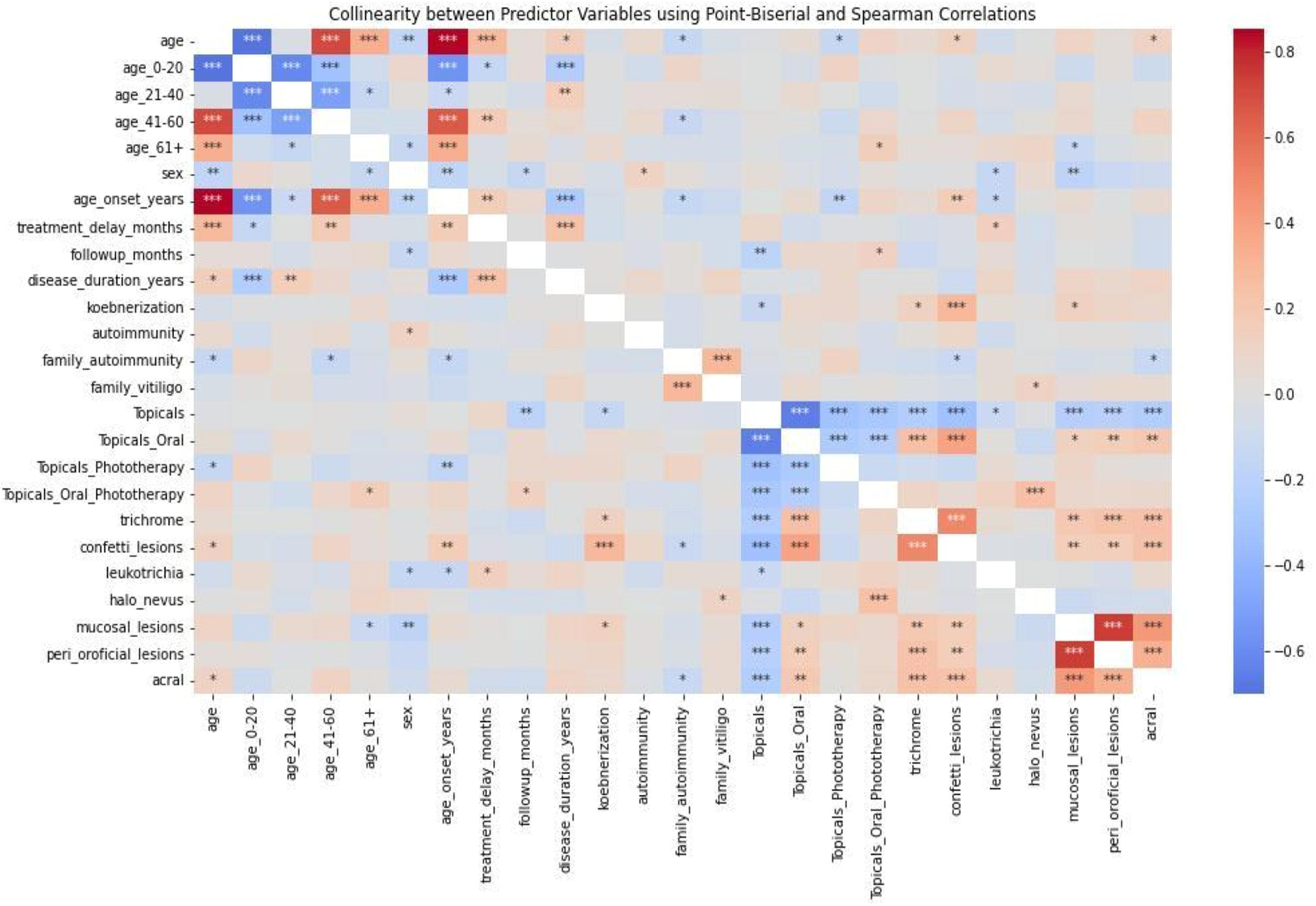
Heatmap showing collinearity among variables. [Dark red indicate strong positive correlations, light red / orange shades represent moderate positive correlations, blue shades indicate negative correlations]

**Table 1.**
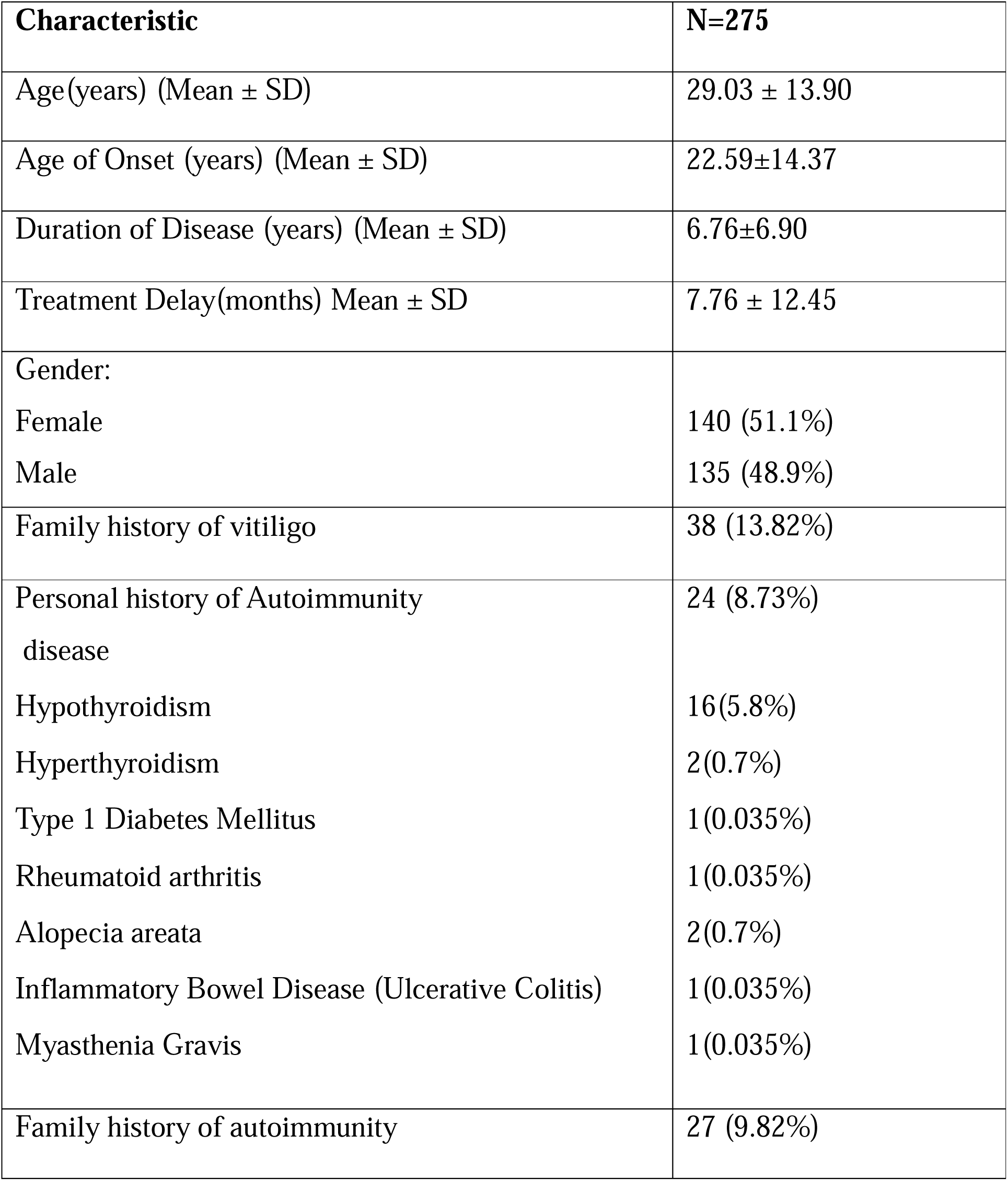
depicts the baseline demographic data.

### Clinicodemographic profile associated with larger BSA involvement

Older age at presentation and later onset of vitiligo were significantly associated with greater baseline BSA involvement (OR 1.03; p=0.046 and p=0.036, respectively). Early-onset vitiligo (0–15 years) showed a protective association (OR 0.47; p=0.047). While personal and familial autoimmunity showed positive but non-significant associations, several clinical markers were robustly predictive of larger BSA. These included koebnerization (OR 2.10; p=0.033), trichrome pattern (OR 3.10; p=0.001), confetti lesions (OR 3.29; p=0.001), leukotrichia (OR 3.69; p<0.001), periorificial lesions (OR 2.09; p=0.043), and acral involvement (OR 2.19; p=0.037). The strength of these associations persisted at 12 months, except periorificial lesions, which did not reach statistical significance (p=0.10).

Correlation analysis showed that increasing BSA across baseline, 6, and 12 months correlated with markers of disease activity (trichrome, confetti, koebnerization), specific site involvement (mucosal, periorificial, acral), and with longer disease duration, later onset, and treatment delay. These trends attenuated over time with treatment. As expected, patients with more extensive disease received aggressive therapies including phototherapy and combination regimens, while limited disease was more often managed with topical modalities (Table 2, Figure 3a, 3b).

**Figure 3:**
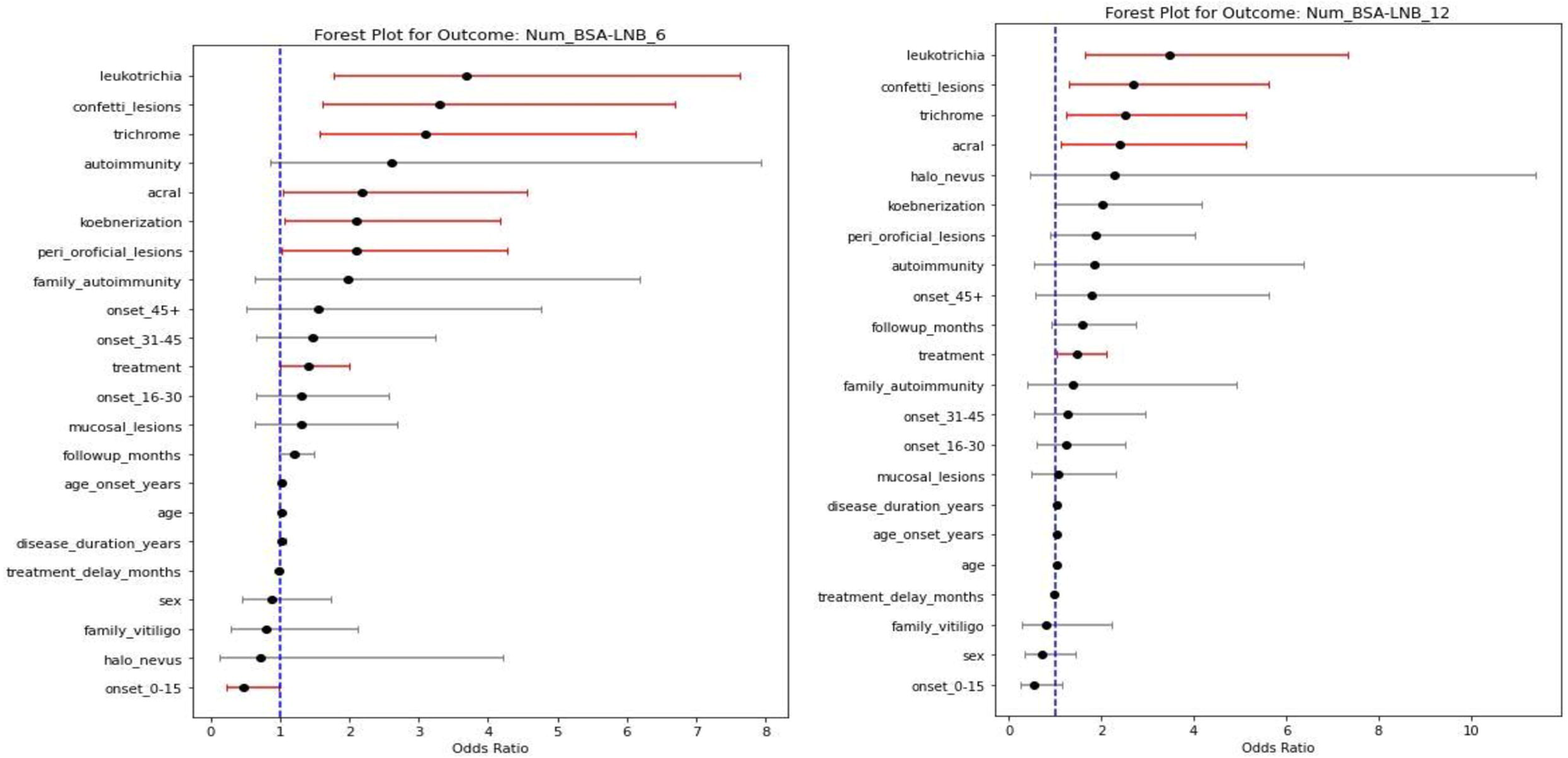
Forest plot displaying the odds ratios (ORs) and 95% confidence intervals of predictors of progression with respect to larger body surface area at 6 months (a) and 12 months (b).

**Table 2.**
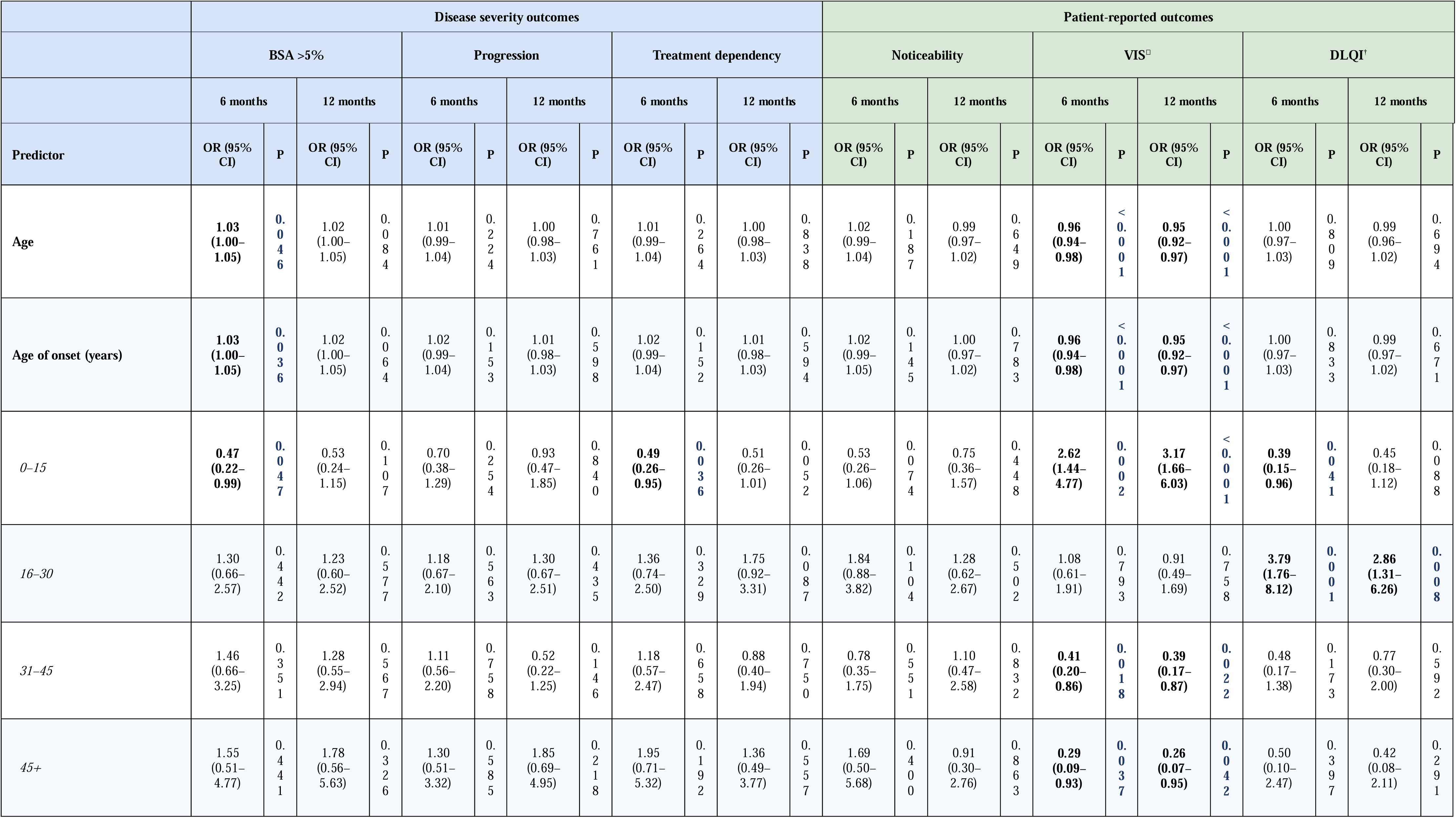
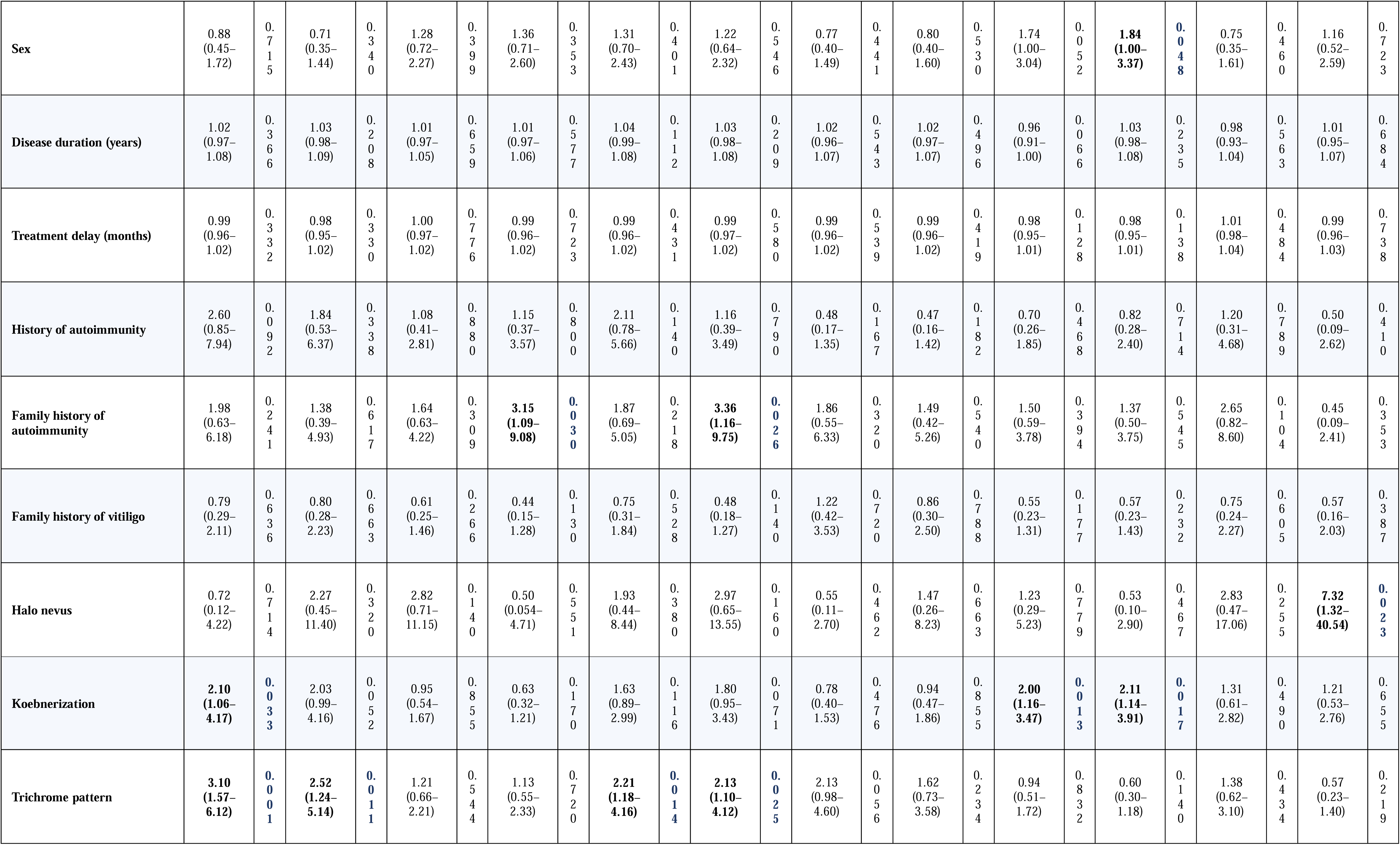
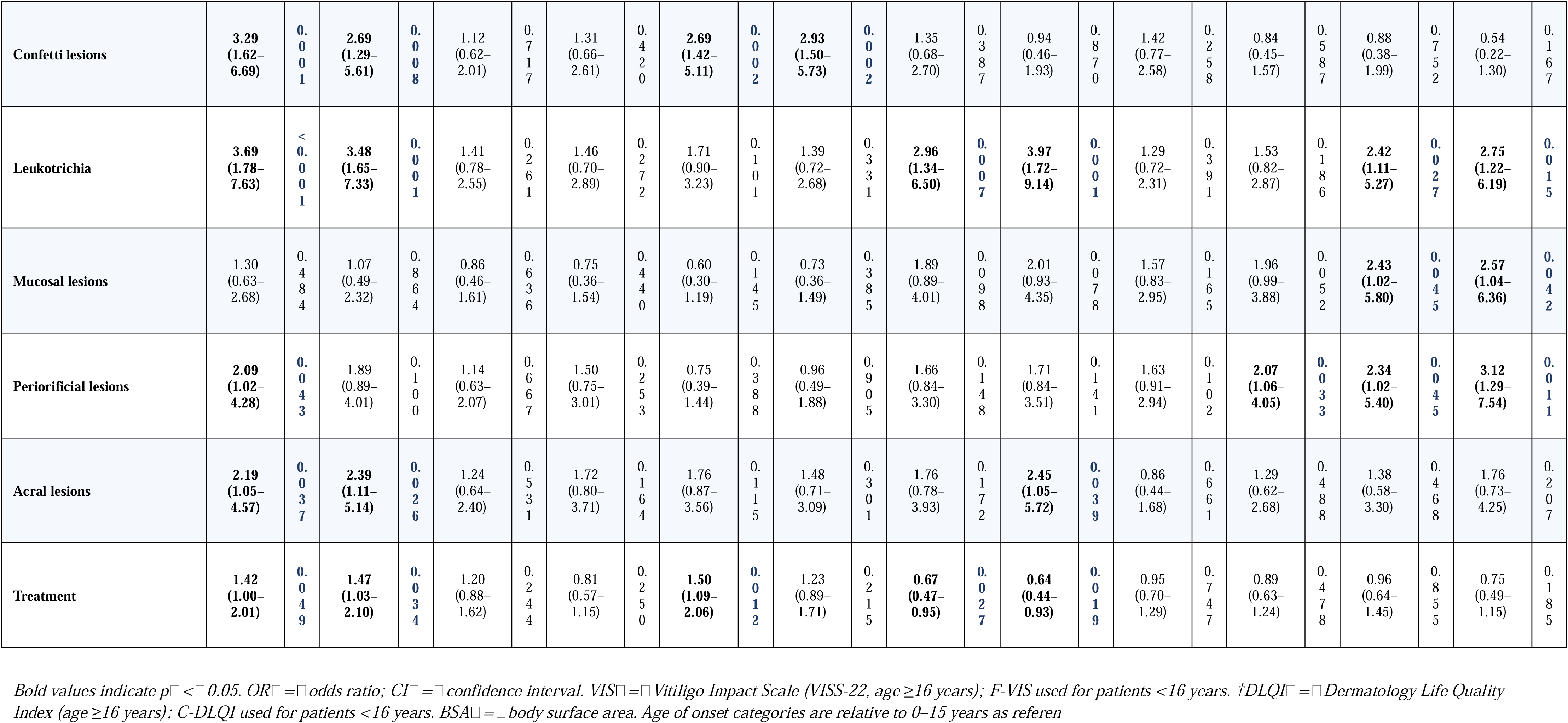
Logistic regression analysis of predictors of disease severity and patient-reported outcomes at 6 and 12 months with OR (95% CI) and p-value.

### Predictors of progression

At 6 months, none of the demographic or clinical factors significantly predicted disease progression. Family history of autoimmunity showed higher—but non-significant—odds of progression (OR 1.64), while family history of vitiligo and early-onset disease showed negative but non-significant associations. By 12 months, family history of autoimmunity emerged as a significant predictor of progression (OR 3.15; p=0.03). No clinical examination features were significantly associated with progression at either time point (Table 2, Figure 4a and 4b).

**Figure 4:**
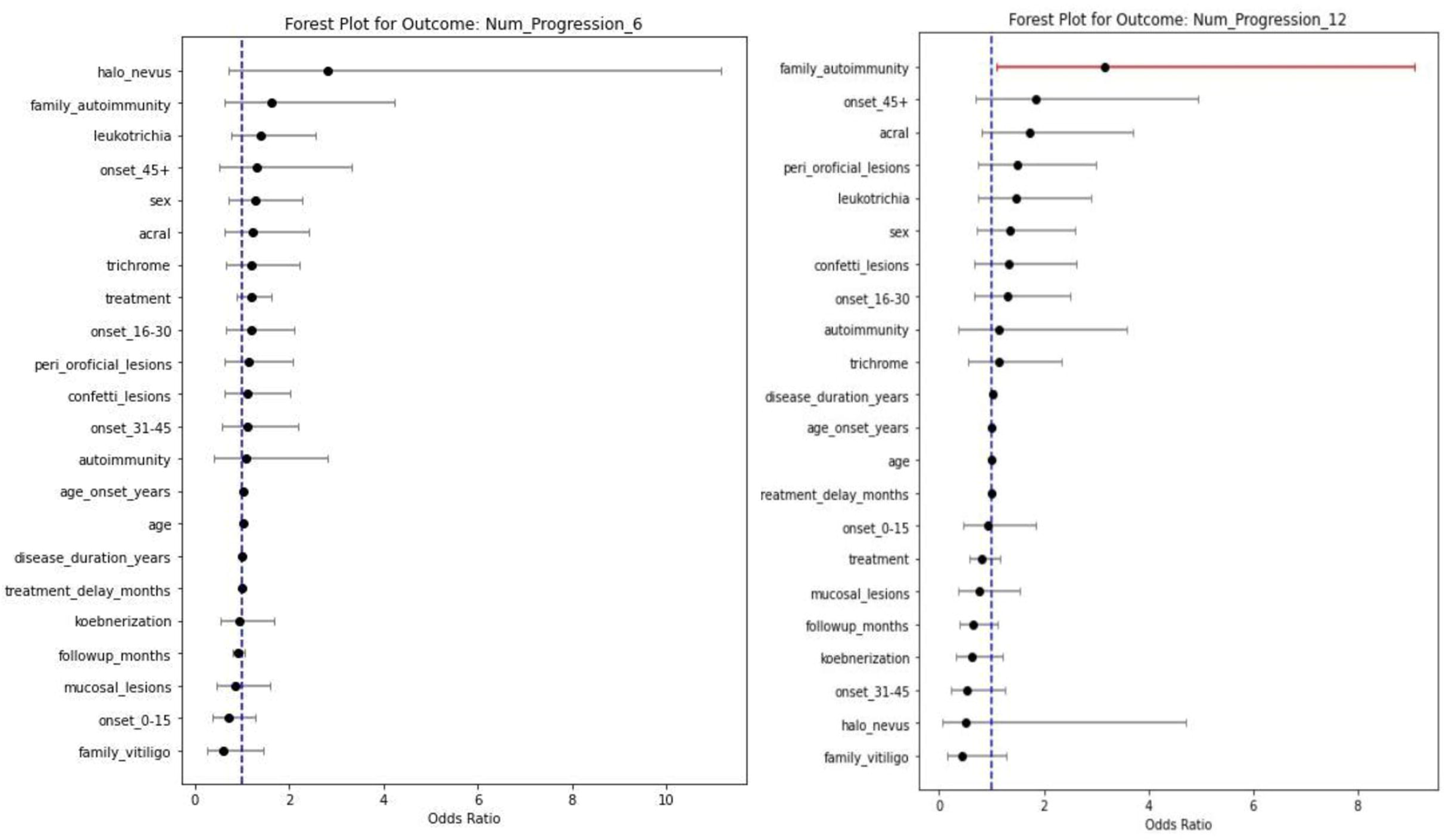
Forest plot displaying the odds ratios (ORs) and 95% confidence intervals of clinical factors predicting progression at 6 months (a) and 12 months (b).

### Predictors of poor repigmentation

At 6 months, early-onset disease (0–15 years) demonstrated a non-significant trend toward better repigmentation (OR 0.58), while family history of vitiligo and autoimmunity demonstrated non-significant poorer outcomes. Among clinical markers, leukotrichia was the only significant predictor of poor repigmentation (OR 2.50; p=0.047). Other markers such as trichrome pattern, mucosal and acral involvement, and periorificial lesions showed positive but non-significant associations with poor repigmentation. Negative predictors included halo nevus and koebnerization, though these were non-significant. At 12 months, this pattern remained largely unchanged, with leukotrichia persisting as the most consistent marker of poor repigmentation (Table 2, and Figure 5a and 5b).

**Figure 5:**
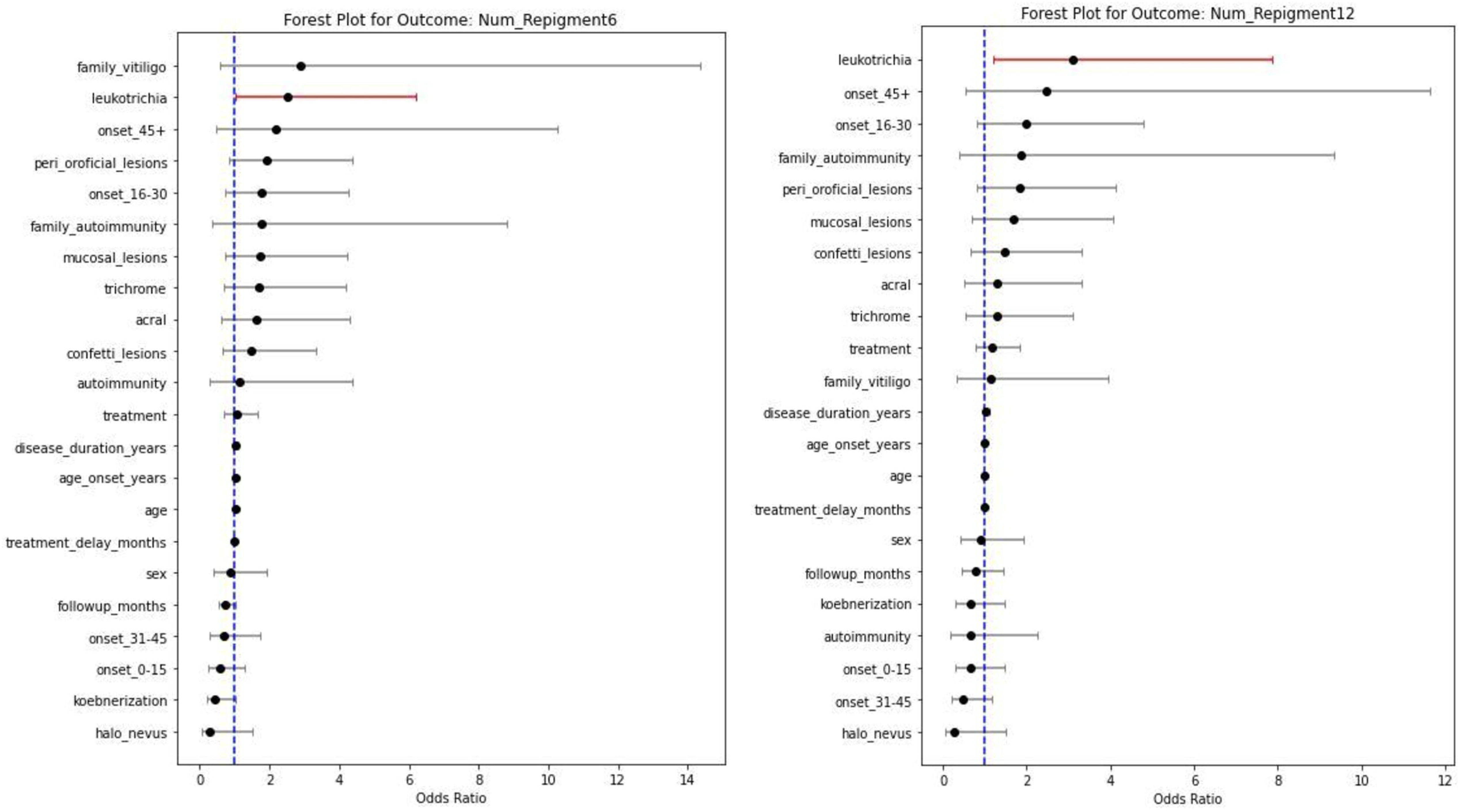
Forest plot displaying the odds ratios (ORs) and 95% confidence intervals of clinical factors predicting poor repigmentation at 6 months (a), at 12 months (b).

### Predictors of treatment dependency

At 6 months, patients with personal or family history of autoimmunity, and those with later age of onset, showed higher but non-significant odds of treatment dependency. Examination findings, however, demonstrated strong associations. Confetti lesions (OR 2.69; p=0.002) and trichrome pattern (OR 2.21; p=0.014) were significantly associated with treatment dependency, along with receipt of combined therapy (OR 1.50; p=0.012). Early-onset disease (0–15 years) was negatively associated with treatment dependency (OR 0.49; p=0.036). At 12 months, confetti lesions (OR 2.93; p=0.002) and trichrome pattern (OR 2.13; p=0.025) remained significant predictors. Mucosal involvement, halo nevus, and koebnerization showed positive but non-significant associations (Table 2, Figure 6a, 6b).

**Figure 6:**
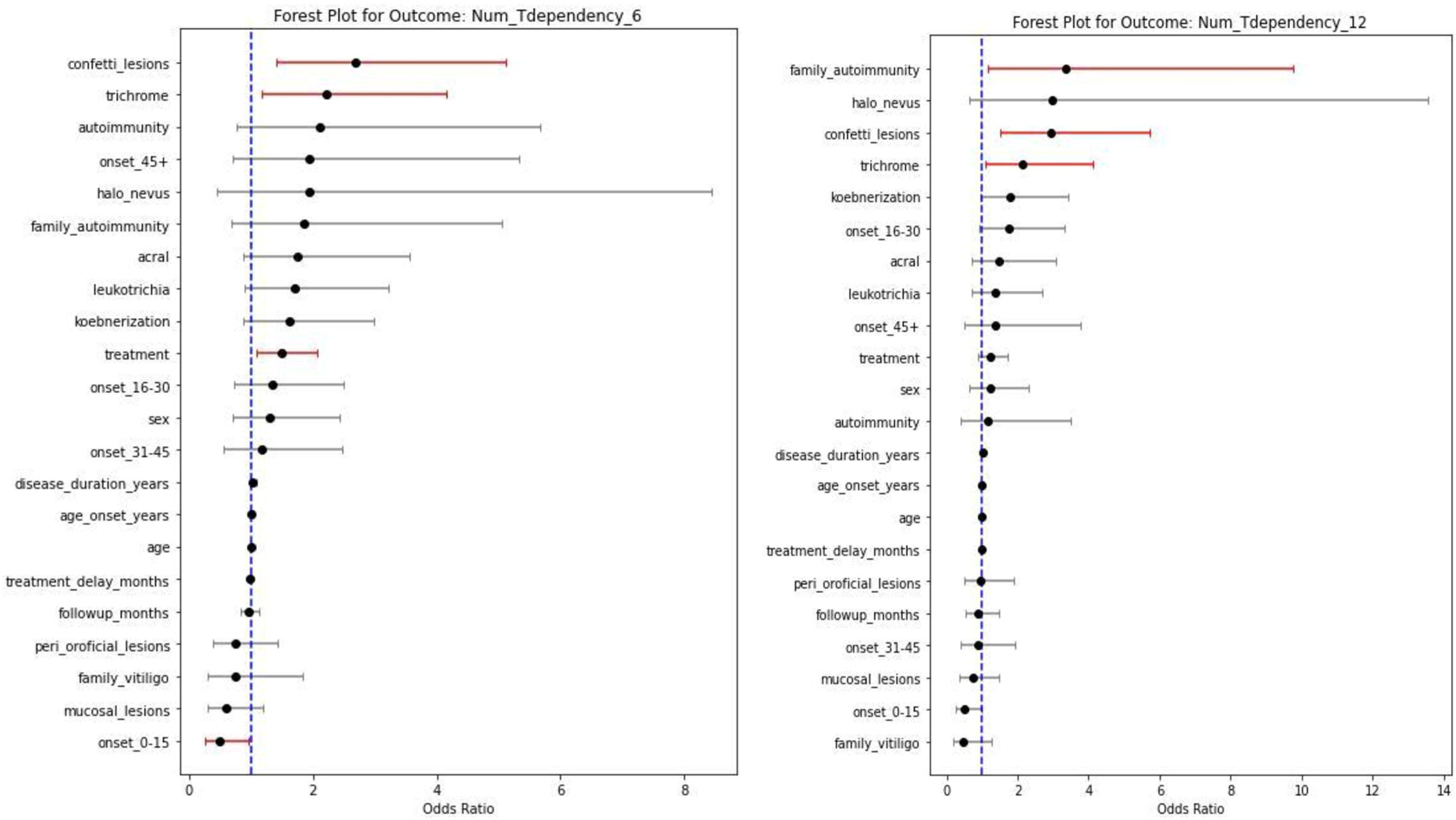
Forest plot displaying the odds ratios (ORs) and 95% confidence intervals of clinical factors determining treatment dependency at 6 months (a) and 12 months (b).

### Predictors of vitiligo noticeability

At 6 months, leukotrichia was significantly associated with higher vitiligo noticeability (OR 2.96; p=0.007); combined treatment showed a significant protective effect (OR 0.67; p=0.027). Clinical markers such as trichrome pattern, mucosal lesions, periorificial lesions, and acral involvement showed positive but non-significant associations. At 12 months, leukotrichia (OR 3.97; p=0.001) and acral lesions (OR 2.45; p=0.039) remained significant predictors of higher VNS. Combined therapy continued to show a significant protective association (OR 0.64; p=0.019). Negative associations were observed with history of autoimmunity and halo nevus. (Table 2, Figure 7a, 7b)

**Figure 7:**
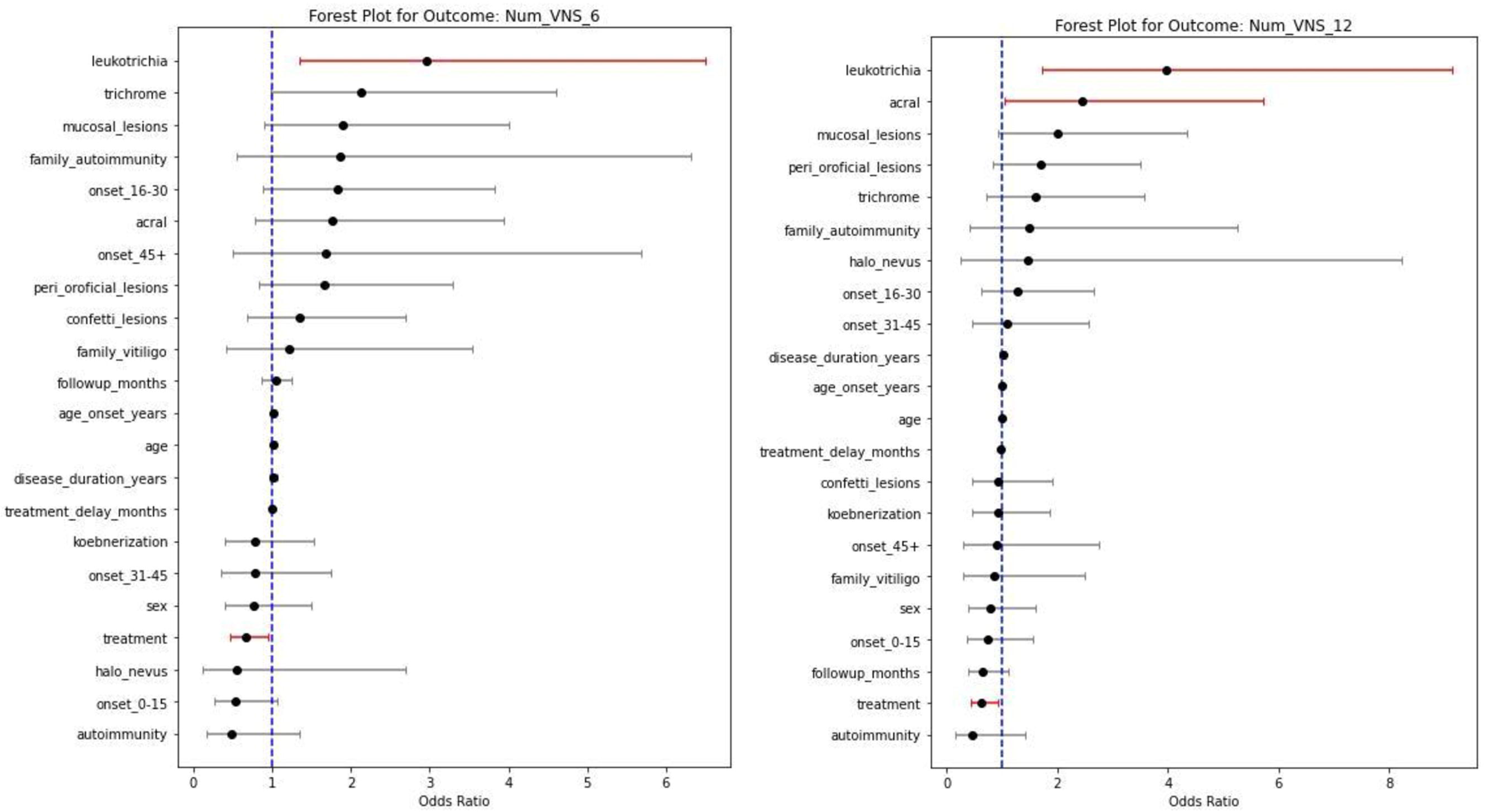
Forest plot displaying the odds ratios (ORs) and 95% confidence intervals of clinical factors associated with vitiligo noticeability at 6 months (a) 12 months (b), Heatmap-Association between factors and continuous outcomes through Phi, Biseriel and spearman coefficients (c)

Continuous VNS analysis showed that later onset, treatment delay, family history of autoimmunity or vitiligo, trichrome pattern, leukotrichia, acral, mucosal and peri-orificial involvement were associated with lower satisfaction scores. Higher VNS, indicating reduced noticeability, was associated with early age of onset, female sex, and use of phototherapy. (Figure 7c)

### Quality of life

#### VISS-22 / F-VIS

Early-onset disease (0–15 years) and koebnerization were significantly associated with higher psychosocial burden in family (measured by F-VIS) at 6 months (OR 2.62; p=0.002 and OR 2.00; p=0.013). Female sex, mucosal and periorificial lesions also showed positive but non-significant associations. At 12 months, early-onset disease remained a strong predictor (OR 3.17; p<0.001), along with koebnerization (OR 2.11; p=0.017), periorificial lesions (OR 2.07; p=0.033) and female sex (OR 1.84; p=0.048). Mucosal lesions and leukotrichia also showed increased but non-significant odds (Table 2, Figure 8a, 8b).

**Figure 8:**
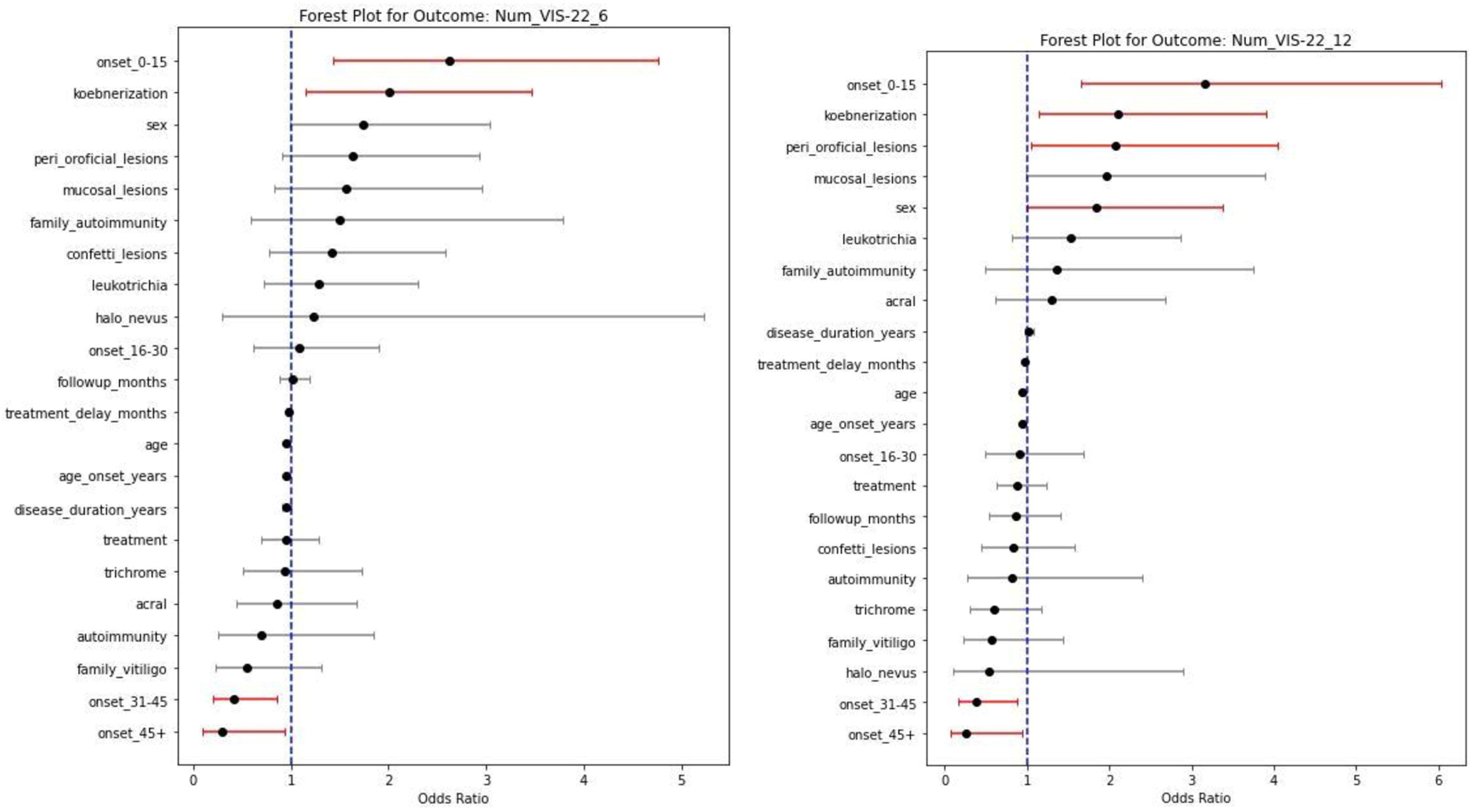
Forest plot displaying the odds ratios (ORs) and 95% confidence intervals of quality of life, (Vitiligo impact scale (VIS)-22= Age ≥16 years, Familial-VIS <16 years) at 6 months (a) 12 months (b)

#### DLQI= Age ≥16 years, C-DLQI <16 years

At 6 months, leukotrichia, mucosal involvement, periorificial lesions, and onset at 16–30 years were significantly associated with poorer quality of life. Early-onset disease (0–15 years) was associated with less psychosocial impact among children(OR 0.39; p=0.041). These associations persisted at 12 months, with halo nevus, leukotrichia, mucosal lesions, and periorificial lesions maintaining significant impact on QoL. Several demographic and clinical variables showed negative but non-significant associations (Table 2, Figure 9a, 9b).

**Figure 9:**
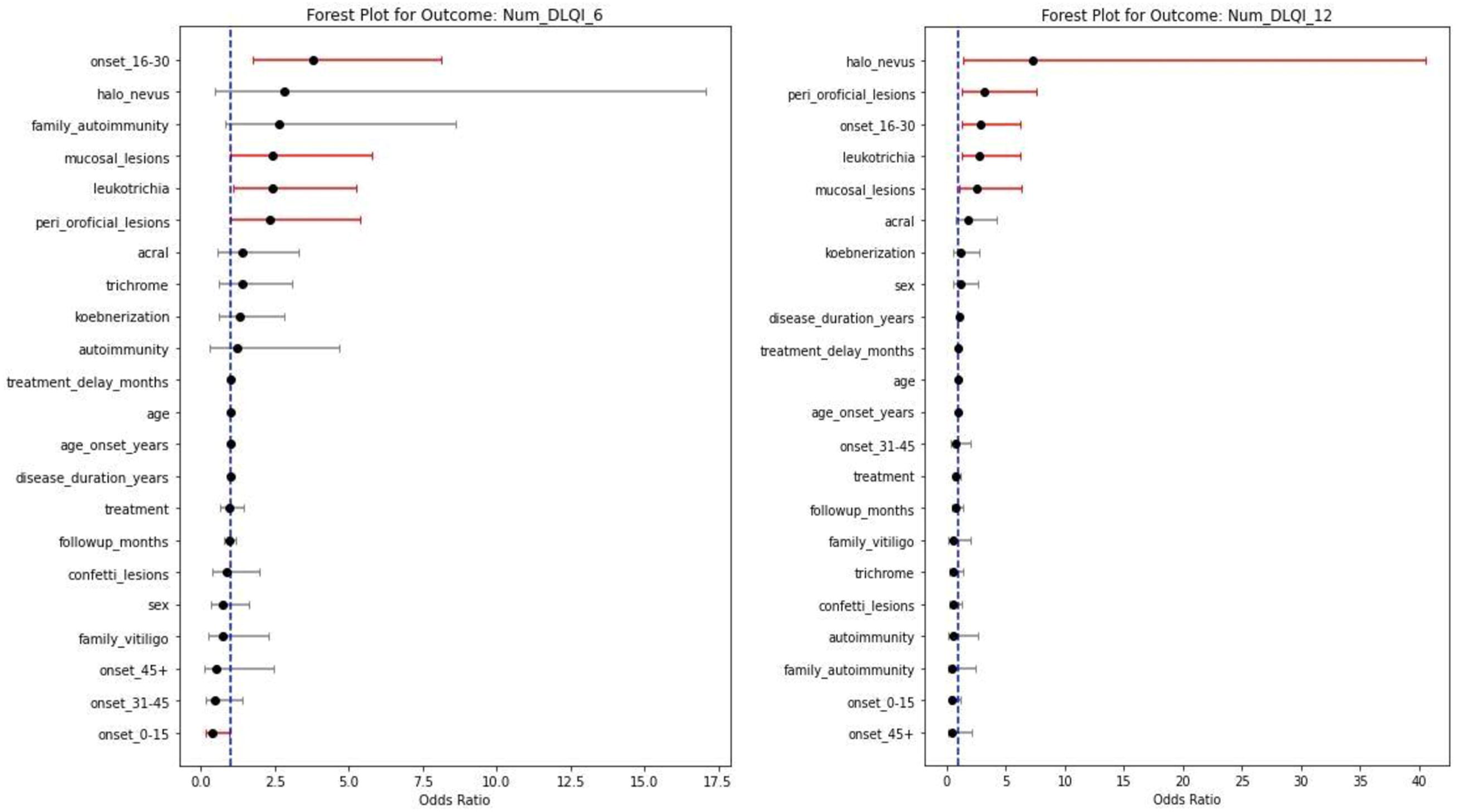
Forest plot displaying the odds ratios (ORs) and 95% confidence intervals of quality of life (dermatology life quality index (DLQI)= Age ≥16 years, childhood-DLQI <16 years) at 6 months (a) 12 months (b)

## Discussion

Non-segmental vitiligo is a chronic, acquired depigmenting disorder characterized by selective melanocyte loss, with a multifactorial pathogenesis involving genetic susceptibility, autoimmunity, oxidative stress, and neurogenic signalling. Despite being a common pigmentary disease, its clinical course is heterogeneous and often unpredictable, making prognostication difficult. A recent scoping review highlighted family history, mucosal involvement, and the presence of clinical activity markers (koebnerization, trichrome, or confetti lesions) as consistent predictors of poor prognosis across studies.^3^

Generalized vitiligo was the predominant subtype (66.18%), similar to earlier studies; however, the proportion of acrofacial vitiligo was notably higher (30.2%), likely reflecting differences in classification thresholds. Leukotrichia (34.55%) resembled previously reported frequencies.^4^ Koebnerization (56.36%) was more prevalent than in prior cross-sectional studies, indicating a higher proportion of active disease at presentation.^5^ Trichrome lesions were found in 34.43% of patients, aligning with available data.^6,7^

Early age of onset (0–15 years) was associated with reduced odds of extensive disease at baseline and 6 months, contrasting with mixed findings in earlier reviews, where some studies reported associations with greater disease extent. Personal and family history of autoimmunity showed positive but non-significant associations, similar to a prior study linking autoimmune comorbidity to >10% BSA involvement. Koebnerization showed a significant or borderline association with extent at follow-up, consistent with multiple prior studies reporting higher BSA involvement in its presence. Trichrome, confetti lesions, leukotrichia, and acral involvement showed the strongest and most consistent associations with larger BSA, concordant with several earlier reports. Periorificial lesions were significant at 6 but not 12 months, reflecting temporal variability. Combination therapy correlated with larger BSA across time points, likely reflecting clinical practice of intensifying treatment in more extensive disease.^8–10^

### Progression

At baseline, confetti lesions, trichrome, and combined treatment were significantly associated with progression. By 6 months, no variable retained significance, possibly due to the effect of early escalation of therapy in high-risk patients. At 12 months, family history of autoimmunity emerged as a significant predictor an association not previously described suggesting a role in long-term disease activity. In contrast to earlier studies identifying clinical activity markers as predictors of progression, our cohort showed no such association at 12 months, possibly reflecting the modifying effect of timely treatment. Early disease onset showed negative, though non-significant, associations with progression, contrary to earlier reports linking childhood onset and family history of vitiligo with progressive disease.^15,11–13^

### Repigmentation

Leukotrichia was a robust independent predictor of poor repigmentation at both 6 and 12 months, with increasing strength over time. This aligns with literature describing leukotrichia as a marker of follicular melanocyte reservoir depletion. Periorificial lesions, mucosal involvement, acral lesions, and clinical markers showed positive but non-significant associations due to poor reservoir and poorer therapeutic response in these areas. Early age of onset, halo nevi, and koebnerization showed non-significant negative associations, reflecting trends toward better repigmentation. Prior studies similarly reported no adverse impact of halo nevi or Koebner’s phenomenon on repigmentation.^15,16,14–17^

### Treatment dependency

Confetti lesions and trichrome pattern showed the strongest associations with treatment dependency across time points, indicating their relevance as markers of a relapsing–remitting course necessitating sustained therapy. Koebnerization approached significance, further supporting its association with chronic activity. Combination therapy showed significant association with dependency, representing both severity-driven treatment selection and genuine therapeutic need.^17–21^ Family history of autoimmunity was significantly associated at 12 months, reinforcing its role in chronicity. Younger age of onset (0–15 years) consistently showed negative associations with dependency/significant at 6 months and borderline at 12 months contradicting earlier studies linking childhood onset and family history to recurrent disease.^21,22^

### Vitiligo noticeability score

Leukotrichia was strongly associated with higher noticeability scores at both time points, reflecting the cosmetic impact of white hair over depigmented patches. Acral lesions were significant at 12 months, consistent with high visibility and resistance to treatment. Mucosal and peri-orificial lesions also showed positive associations, though non-significant. Combination therapy was associated with reduced noticeability, highlighting improved cosmetic outcomes. Previous surveys similarly reported better satisfaction with phototherapy over topical therapy.^23–25^

### Quality of life

Early-onset vitiligo (0–15 years) was associated with greater family burden, reflected by higher Family-VIS scores, indicating parental distress. In contrast, young children had relatively low DLQI, whereas onset between 16 and 30 years was linked to higher DLQI, likely due to concerns related to education, employment, relationships, and marriage. Female sex was strongly associated with greater psychosocial impact, consistent with prior literature. Koebnerization, periorificial and mucosal involvement, and leukotrichia were also linked to higher psychosocial burden. Older age at onset and a family history of vitiligo showed inverse associations, possibly reflecting better coping and disease familiarity.^25–27^

### Limitations

Study was conducted at a tertiary care center, a setting where patients with more severe and refractory disease are registered, which may limit the generalizability of our findings to the broader population and study period of one year is short for a chronic relapsing disease like vitiligo.

## Conclusion

Overall, we found leukotrichia, trichrome and confetti patterns, koebnerization, and mucosal or periorificial involvement indicate poorer prognosis, greater disease extent, and reduced repigmentation. Family history of autoimmunity predicts progression and treatment dependency. Early-onset cases show lower extent but higher psychosocial burden. Incorporating these markers supports early risk stratification, individualized therapy, and improved outcomes. Furthermore, efforts to develop the proposed scoring system and to undertake its external validation across diverse ethnic and geographic cohorts is currently ongoing.

## Data Availability

All data produced in the present study are available upon reasonable request to the authors

---

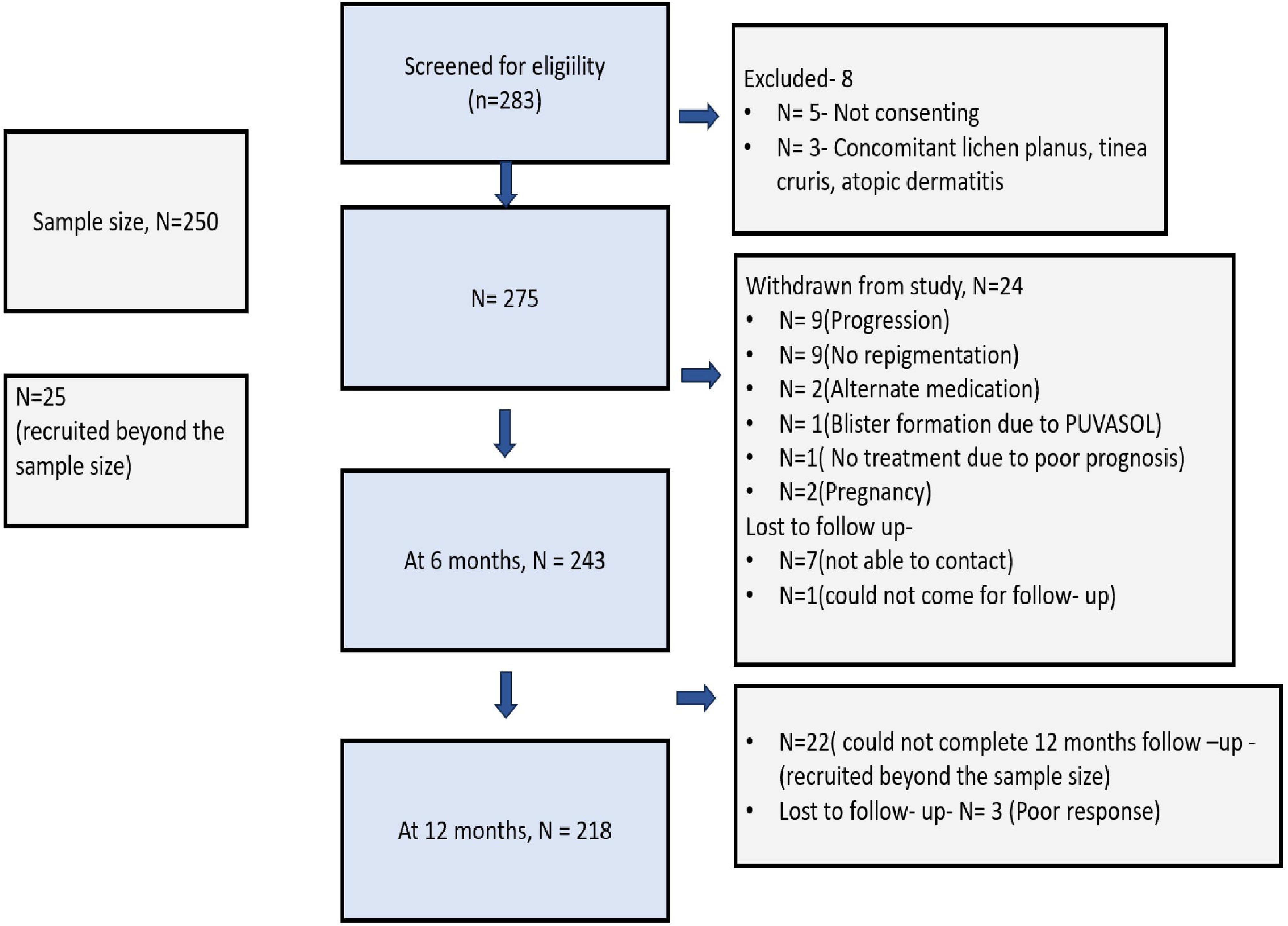

